# Mental health priorities and challenges in Zambia: A scoping study

**DOI:** 10.1101/2025.09.25.25336232

**Authors:** Narinder Bansal, Petros I Andreadis, Philip Chimponda, Zambia Sandra Barteit, Sashi P Sashidharan, Ravi Paul

## Abstract

**Background:** The design and delivery of safe and effective mental healthcare requires data on local needs and priorities. The aim of this scoping review is to provide background information on the prevalence of mental health conditions and local stakeholder experiences of mental healthcare in Zambia.

**Methods:** We searched electronic databases of published (Medline, PsycINFO, Embase, African Index Medicus) and unpublished (University of Zambia repository) literature to retrieve relevant epidemiological and qualitative articles from database inception to January 9^th^, 2024. Qualitative studies were synthesised using thematic synthesis and key themes were triangulated with experiences of local stakeholders.

**Results:** Eleven epidemiological papers were identified. These reported on the prevalence of mental distress in the general population (16.9%); depressive symptoms in adolescents (29.7%); problematic alcohol consumption in the general population (dependence, 7.4%; binge, 11.6%; and unhealthy consumption, 15.3%) and in adolescents (45.1%); suicidal ideation (7.8%) and behaviour (8.5%) in the general population and in adolescents (31.3% and 39.6%, respectively); suicide attempts in the general population (2.3%). Synthesis of 10 qualitative articles identified interrelated themes relating to barriers to access and provision of mental healthcare. Mental health stigma is perceived to be pervasive across all sectors of society and partly attributed to the language used in the previous Mental Health Act and the national psychiatric hospital. Structural stigma is perceived to drive the low priority of mental health in Zambia in policy, funding, advocacy and research. Reported consequences include low availability of safe and effective mental healthcare, particularly at community level, resulting in a cycle of coercive hospital admission, discharge, relapse and readmission. This is perceived to place significant social, emotional and economic stress on patients and their families. Carer burnout and the lack of visible recovery perpetuates the stigma that people with mental illness have little value to society.

**Conclusions:** Findings from this review indicate the need for a multisectoral approach to tackle structural stigma, increase national advocacy for mental health, and facilitate the provision of safe and effective community-based mental healthcare in Zambia. While epidemiological data is limited, the current evidence indicates that adolescents are a high priority group for early intervention.

## Background

The design and delivery of safe, effective, and meaningful mental health support and care is a global challenge. Mental health systems all over the world are characterised by significant gaps relating to care, treatment, research, governance, service structures, financial and human resources.(1) These gaps are particularly striking in low-income settings in the Global South where it has been estimated that less than ten percent of people who need treatment for mental illness will receive it.(2) Resource constraints in these settings are compounded by the dominance of institutionalised mental healthcare and a lack of culturally and socially aligned treatment models.(1)

Community uptake of mental health support requires the availability of treatment interventions and pathways that are aligned with local experiences, values, needs and priorities.(3) This is a challenge in many low-income regions, particularly in Sub-Saharan Africa, where there are insufficient resources and capacity to generate, publish, and disseminate local data.(4) A review of epidemiological studies in 2021 found significant research gaps on the prevalence of psychiatric disorders in Africa, with more than half the surveys originating from five countries (Ethiopia, Kenya, Nigeria, South Africa, and Uganda).(5) Other important knowledge gaps include data on the perceptions, experiences, and priorities of local stakeholders across policy, practice, and community settings, including people with lived experience (PWLE) of mental illness. These data are needed to inform local policy and practice and increase equity in global mental health.

In Zambia, the government has been working to align mental health policy and practice with human-rights based approaches in response to local service user reports of systemic abuse and neglect of people experiencing mental illness.(6, 7) This effort includes the revision of the Mental Health Act in 2019 and the creation of a mental health policy in 2005 advocating for the provision of non-coercive community-based mental healthcare. While these changes have been viewed positively by local stakeholders, little progress has been made to bring the aspirations of the legislation and policy to fruition, and centralised hospital-based care remains the main source of formal treatment.(8) Qualitative studies have highlighted some of the obstacles to progressing mental healthcare in Zambia, including the lack of local data on the prevalence of mental illness.(9) However, these studies have not been reviewed to give a nuanced understanding of key challenges and priorities from the perspective of a wide range of local stakeholders. This includes understanding the local impact of the COVID-19 pandemic, which is estimated to have increased the global prevalence of anxiety and depressive disorders by 26% and 28%, respectively.(10)

The aim of this scoping review is to provide background information on the prevalence of mental health conditions and the perceptions and experiences of local stakeholders on mental healthcare in Zambia. Specifically, we sought to identify the key mental health priorities, challenges, and data gaps in terms of illness burden, and access to, and provision of, statutory (provided by the state) mental healthcare in Zambia.

## Methods

### Study design and rationale

We conducted a scoping review of published and grey epidemiological and qualitative literature on mental health in Zambia in line with Preferred Reporting Items for Systematic Reviews and Meta-Analyses for Scoping Reviews (PRISMA-ScR) guidelines.(11) A scoping review was deemed appropriate given the uncertainty about the quantity and quality of available data in Zambia. Scoping review findings were triangulated with experiences of local stakeholders gathered during stakeholder consultation meetings in Zambia (described in the section below ‘*Local stakeholder involvement and triangulation*’).

### Search criteria and strategy

For the epidemiological review (Appendix 1), we identified broad search terms (including free text and subject headings) for ‘mental disorders’ and limited the search to epidemiological studies (study design) in Zambia (population). For the qualitative review (Appendix 2), we identified broad search terms for ‘mental ill health’ (phenomena of interest), ‘help-seeking/’experience” (study focus) and limited the search to qualitative studies in Zambia. We conducted a comprehensive search of electronic databases of published (Medline, PsycINFO, Embase, African Index Medicus) and unpublished literature (University of Zambia repository) from database inception to January 9^th^, 2024. We used more crude search terms for the databases African Index Medicus (mental AND Zambia) and University of Zambia repository (‘mental’, ‘depression’, ‘anxiety’) due to the lack of advanced filters in these databases and found that the search term ‘mental’ identified a wide range of articles relating to distress, common mental disorders, substance use, and severe mental illness. We also consulted local academics and mental health experts to help identify relevant studies and reports. Articles identified by our search strategy were screened against the inclusion and exclusion criteria.

### Inclusion and exclusion criteria

#### Epidemiological literature

We included articles reporting epidemiological data on the incidence or prevalence of mental disorders in Zambia from studies that used census or probabilistic procedures to obtain a national or regional sample. We excluded papers reporting epidemiological studies using convenience population sampling methods, those targeting specific groups (i.e. patient populations) and those with a sample size <400.

#### Qualitative literature

We included articles reporting qualitative studies (using qualitative methods of data collection and analysis) exploring perceptions and experiences of mental health support in Zambia including help-seeking pathways and service use/provision. We excluded qualitative studies that did not use qualitative methods of data collection and analysis and those that focused primarily on exploring determinants of mental illness in Zambia.

### Article screening

Articles identified by the search strategy were title-screened for relevance by NB. Titles identified as relevant were abstract-screened by NB (10% independently double-screened by PA). Abstracts identified for inclusion were full-text screened against the inclusion and exclusion criteria by NB and PA (100% independently double screened).

### Extraction

Key characteristics of included epidemiological and qualitative articles were extracted and tabulated. There were insufficient epidemiological studies to allow synthesis by outcome.

### Qualitative Evidence Synthesis

Following recommendations from previous research,(12) we decided to carry out a thematic synthesis after an initial review of the qualitative papers to assess data richness and suitability.(13) A thematic synthesis was chosen to address our two research questions that seek to explore the perceptions and experiences of a wide range of stakeholders on healthcare: 1.‘*What are local stakeholder perceptions and experiences of mental healthcare in Zambia?’* and 2.*‘What does the qualitative literature reveal in terms of major challenges and priorities in mental healthcare?*’. Thematic synthesis is also suitable for the type of studies included in our review – a mixture of conceptually thin and ‘thick’ data.(13) Each paper was critically read and reread by two researchers (NB and PA). Papers were coded line by line following an iterative process of identifying codes and developing a coding list relevant to the research question. We identified descriptive themes (themes close to the primary data) and more analytical themes (new interpretative constructs). We identified, conceptualised and mapped out the relationship between these themes.

### Local stakeholder involvement and triangulation

We consulted fifteen stakeholders from Zambia throughout the review process to discuss emerging and final findings. These stakeholders provided lived experience perspectives and experiences from diverse lay and professional backgrounds (including community, statutory mental health care, medical education, third sector mental health, private sector mental health, primary and secondary education). Stakeholder feedback helped provide data triangulation and determine the extent to which extracted themes were still relevant.

## Results

### Selection of studies

#### Epidemiological literature

As shown in Figure 1, our search strategy for epidemiological literature identified 1630 articles. These articles were title-screened for relevance by NB resulting in 99 identified as relevant (reporting quantitative data relating to mental health in Zambia). These titles were abstract screened for relevance (10% double screened by PA with 100% agreement) resulting in 22 articles identified for full-text screening. Full texts of these articles were double-screened (independently by NB and PA) against the inclusion and exclusion criteria, with 11 articles meeting these criteria.

**Figure 1:**
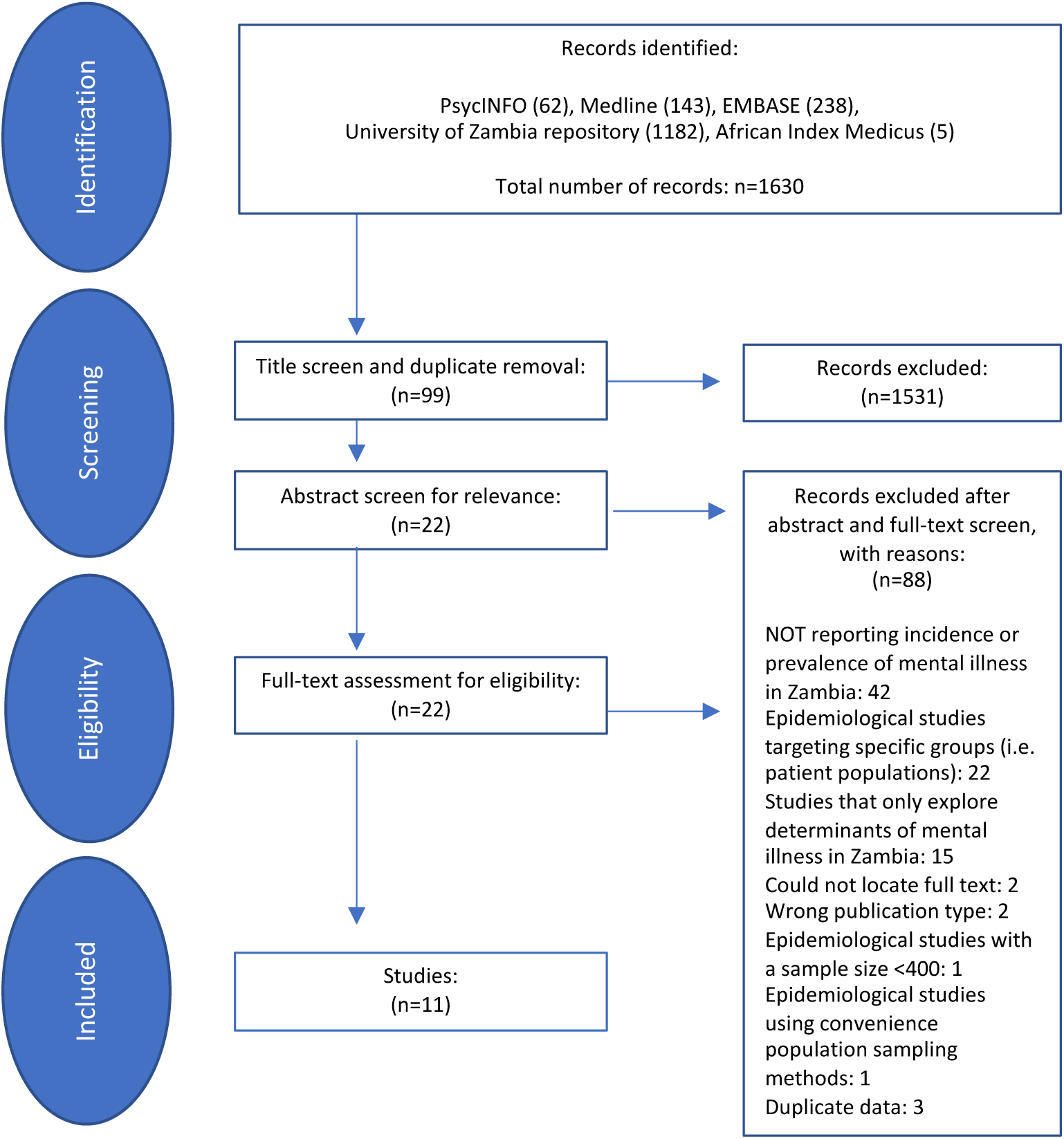
Outcome of study selection (epidemiological literature)

#### Qualitative literature

As shown in Figure 2, our search strategy for qualitative literature identified 2181 articles. These articles were title screened for relevance by NB, resulting in 22 identified as relevant (reporting qualitative data relating to mental health in Zambia). These titles were abstract screened for relevance (three double screened by PA with 100% agreement) resulting in 11 articles identified for full-text screen. Full texts of these articles were double-screened (independently by NB and PA) against the inclusion and exclusion criteria and 10 articles were identified as meeting these criteria.

**Figure 2:**
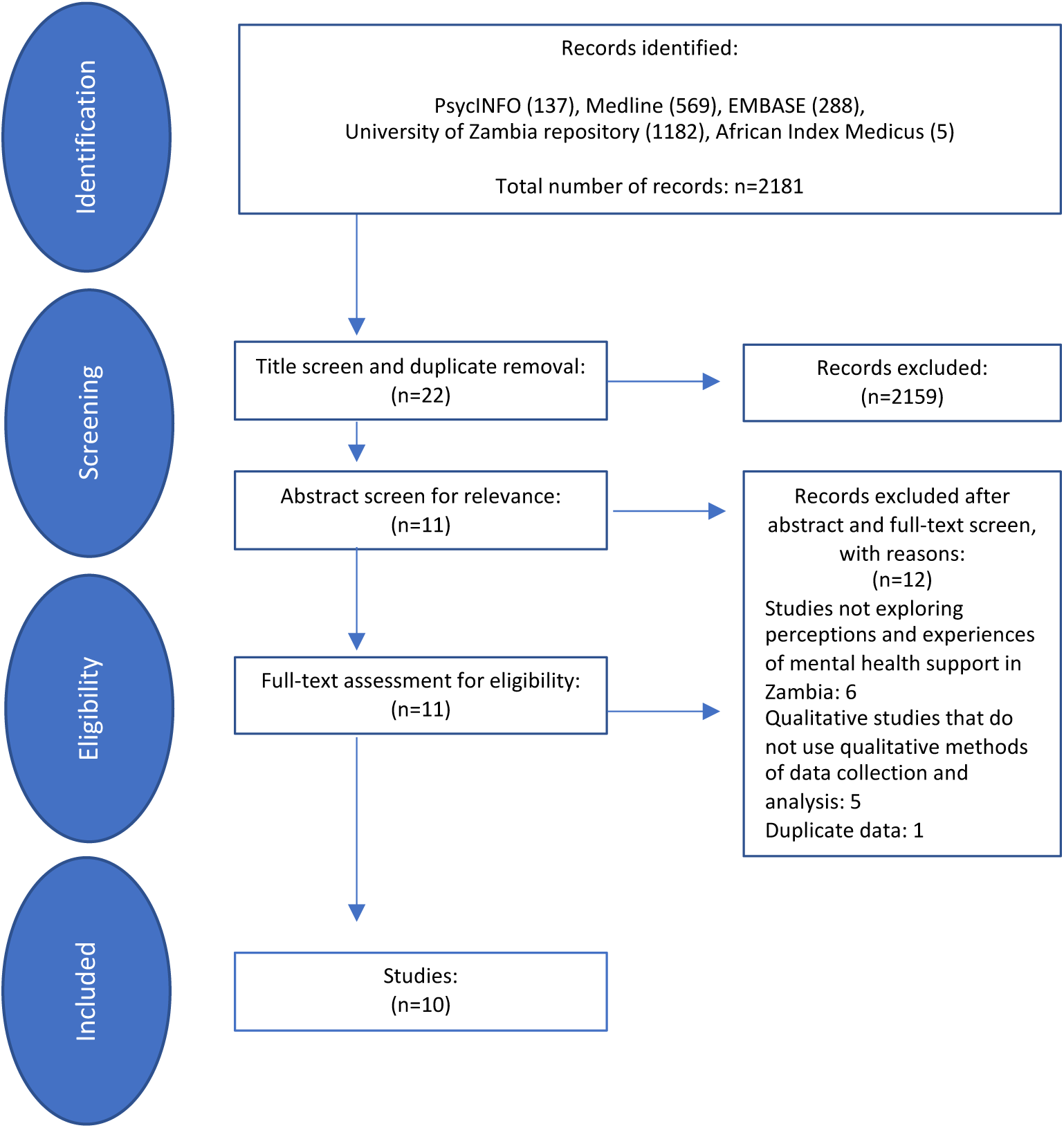
Outcome of study selection (qualitative literature)

### Characteristics of included epidemiological studies

We identified 11 papers reporting on the prevalence of mental health conditions in Zambia including mental distress, harmful alcohol consumption, self-harm, and suicidal ideation/behaviour. Table 1 shows characteristics of these 11 epidemiological papers, listed by mental health condition. All prevalence estimates were measured using self-reporting questionnaires. Seven papers reported data from two global epidemiological surveys: five papers report data from the 2004 Global School-Based Student Health Survey of students (GSHS) and two papers report data from the World Health Organization (WHO) STEPwise approach to non-communicable diseases risk factor surveillance (STEPS) survey. With the exception of one study (14), all papers reported data from studies conducted prior to the COVID-19 pandemic. A large proportion of the included papers focus on adolescent health (n=6), with sample size ranging from 1,241 to 20,923 participants.

**Table 1:**
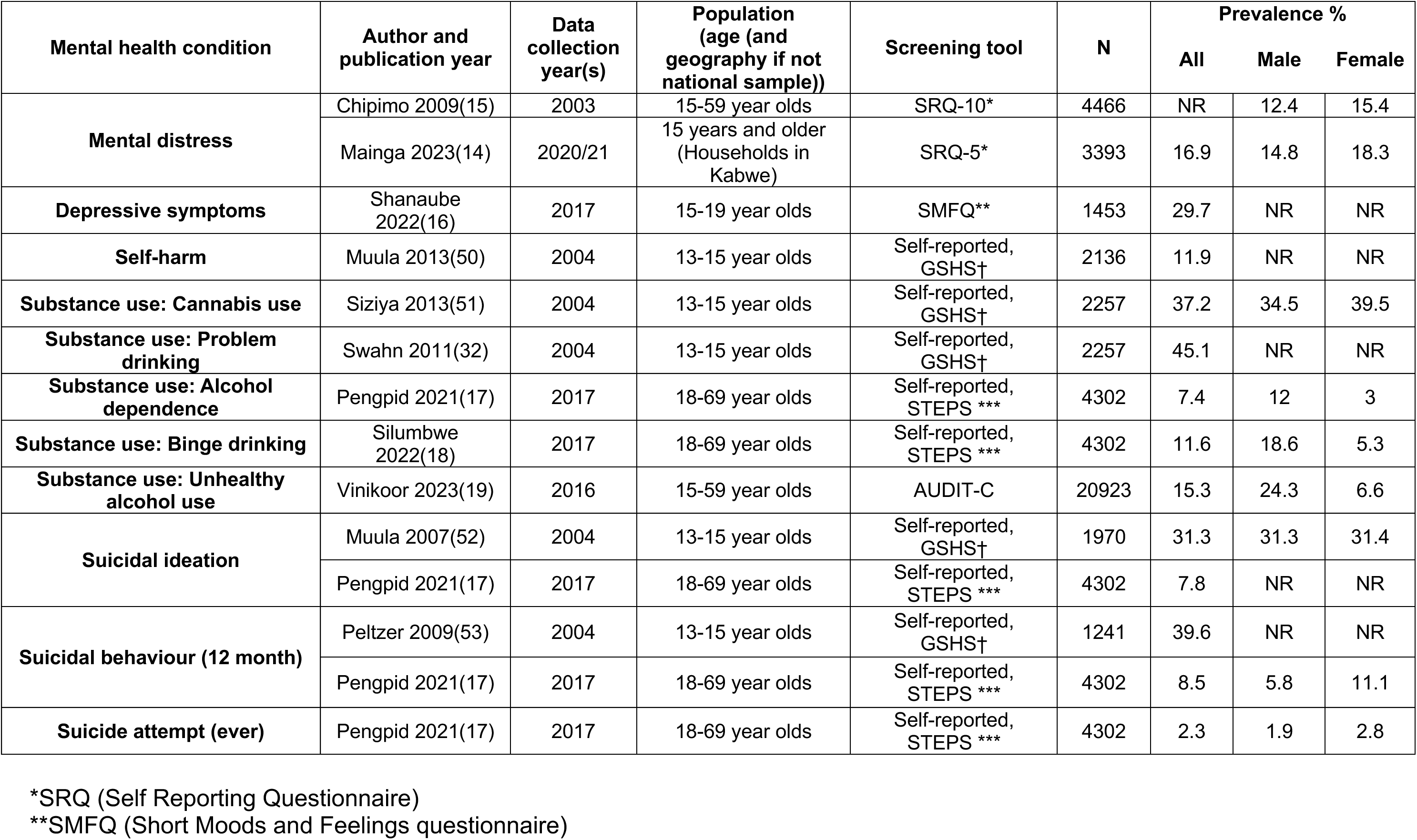
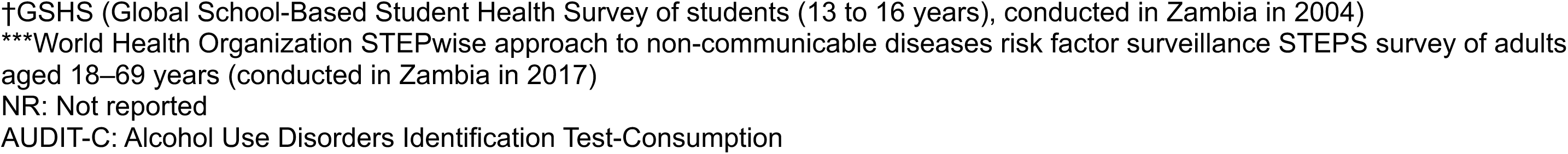
Details of included studies by mental health condition: (epidemiological literature)

| Mental health condition | Author and publication year | Data collection year(s) | Population (age (and geography if not national sample)) | Screening tool | N | Prevalence % |  |  |
| --- | --- | --- | --- | --- | --- | --- | --- | --- |
|  |  |  |  |  |  | All | Male | Female |
| Mental distress | Chipimo 2009(15) | 2003 | 15-59 year olds | SRQ-10* | 4466 | NR | 12.4 | 15.4 |
|  | Mainga 2023(14) | 2020/21 | 15 years and older (Households in Kabwe) | SRQ-5* | 3393 | 16.9 | 14.8 | 18.3 |
| Depressive symptoms | Shanaube 2022(16) | 2017 | 15-19 year olds | SMFQ** | 1453 | 29.7 | NR | NR |
| Self-harm | Muula 2013(50) | 2004 | 13-15 year olds | Self-reported, GSHS† | 2136 | 11.9 | NR | NR |
| Substance use: Cannabis use | Siziya 2013(51) | 2004 | 13-15 year olds | Self-reported, GSHS† | 2257 | 37.2 | 34.5 | 39.5 |
| Substance use: Problem drinking | Swahn 2011(32) | 2004 | 13-15 year olds | Self-reported, GSHS† | 2257 | 45.1 | NR | NR |
| Substance use: Alcohol dependence | Pengpid 2021(17) | 2017 | 18-69 year olds | Self-reported, STEPS *** | 4302 | 7.4 | 12 | 3 |
| Substance use: Binge drinking | Silumbwe 2022(18) | 2017 | 18-69 year olds | Self-reported, STEPS *** | 4302 | 11.6 | 18.6 | 5.3 |
| Substance use: Unhealthy alcohol use | Vinikoor 2023(19) | 2016 | 15-59 year olds | AUDIT-C | 20923 | 15.3 | 24.3 | 6.6 |
| Suicidal ideation | Muula 2007(52) | 2004 | 13-15 year olds | Self-reported, GSHS† | 1970 | 31.3 | 31.3 | 31.4 |
|  | Pengpid 2021(17) | 2017 | 18-69 year olds | Self-reported, STEPS *** | 4302 | 7.8 | NR | NR |
| Suicidal behaviour (12 month) | Peltzer 2009(53) | 2004 | 13-15 year olds | Self-reported, GSHS† | 1241 | 39.6 | NR | NR |
|  | Pengpid 2021(17) | 2017 | 18-69 year olds | Self-reported, STEPS *** | 4302 | 8.5 | 5.8 | 11.1 |
| Suicide attempt (ever) | Pengpid 2021(17) | 2017 | 18-69 year olds | Self-reported, STEPS *** | 4302 | 2.3 | 1.9 | 2.8 |

\*SRQ (Self Reporting Questionnaire)
\*\*SMFQ (Short Moods and Feelings questionnaire)
†GSHS (Global School-Based Student Health Survey of students (13 to 16 years), conducted in Zambia in 2004)
\*\*\*World Health Organization STEPwise approach to non-communicable diseases risk factor surveillance STEPS survey of adults aged 18–69 years (conducted in Zambia in 2017)
NR: Not reported
AUDIT-C: Alcohol Use Disorders Identification Test-Consumption

### Prevalence of mental health conditions in Zambia

The prevalence of mental distress in the general population was reported at 16.9% during the COVID-19 pandemic (18 November 2020 to 24 February 2021).(14) Two studies conducted 17 years apart indicate a higher prevalence of mental distress in women compared to men and an increase in prevalence in both genders over time.(14, 15) A study conducted in 2017 indicates that nearly a third of adolescents have depressive symptoms.(16) Data from the 2004 GSHS school surveys indicate a high prevalence of problematic substance use (37.2% reporting cannabis use, 45.1% reporting problematic drinking) and suicidal behaviours (31.3% reporting suicidal ideation, 39.6% reporting suicidal behaviour in the past 12 months) in Zambian adolescents. As shown in Table 1, these estimates are significantly higher than those reported in the general adult population, particularly in the STEPS survey although these data were collected 13 years later. Studies in adults indicate a higher prevalence of problematic alcohol use in men and a higher prevalence of suicidal behaviour in women.(17–19)

### Characteristics of included qualitative studies

We identified 10 papers reporting on the perceptions and experiences of mental healthcare in Zambia. Table 2 shows characteristics of the included qualitative studies. Most studies were conducted prior to the publication of the new Mental Health Act in 2019 and before the onset of the COVID-19 pandemic (n=7). Study participants included a wide range of stakeholders, including service users, community members, health service providers, clinical staff, policy makers, traditional healers, non-governmental organisations (NGO), representatives, educators, police, and academics.

**Table 2:** Details of included studies (qualitative literature)

| **Author and publication year** | **Data collection year** | **Population** | **Number of participants** | **Method of data collection** |
| --- | --- | --- | --- | --- |
| Sichimba 2022(24) | 2021 | Primary caregivers of patients with mental illness treated at Chainama psychiatric hospital | 15 | Interviews |
| Mainga 2022(20) | NR | TB stakeholders and health workers | 162 | Interviews and focus groups |
| Davids 2019(25) | 2017 | CAMH researchers and clinicians | 17 | Interview survey |
| Munakampe 2016(22) | 2016 | Primary caregivers, mental health nurses and doctors, policymakers in three provinces | 12 | Interviews |
| Chilema 2015(27) | NR | Primary health care providers | 20 | Focus groups |
| Karban 2013(26) | NR | Mental health educators and practitioners | 13 | Focus groups |
| Bird 2011(9) | NR | health and mental health policy makers in the Ministry of Health, politicians, health service managers, media representatives, representatives from non-health sectors, health professional unions, traditional healer unions, mental health user groups and mental health non-governmental organizations (NGOs) | 65 | Interviews |
| Mwanza 2011(23) | 2005/2006 | policy makers in and outside of the ministry of health, general and mental health care professionals, users of psychiatric services, teachers, police officers, academics, members of three NGOs and traditional healers | 65 | Interviews and focus groups |
| Kapungwe<br>2010(21) | 2005/2006 | policy makers (from the Ministry of Health and elsewhere), health and mental health care professionals, users of psychiatric services, teachers, police officers, academics, members of three NGOs and traditional healers | 65 | Interviews and focus groups |
| Sikwese(28) | NR | heads of departments of line ministries and units, mental health coordinators, health service managers, lead nurses and clinicians and notable non-governmental organizations (NGOs) dealing with mental illnesses. policy makers in the ministry of health and other ministries, health and mental health care professionals, users of psychiatric services, teachers, police officers, academics, members of three NGOs and traditional healers | 65 | Interviews and focus groups |

### Key themes from thematic synthesis of qualitative data

We identified 16 analytical themes relevant to our research questions, categorised as barriers to mental healthcare (Table 3) and desired solutions and interventions (Table 4). These themes are discussed in detail below. Tables 3 and 4 show how these analytical themes relate to descriptive themes and provide a summary of each theme, using the language and key phrases of the study authors and participants where possible. As shown in Figure 3, the themes related to barriers to mental healthcare were found to be conceptually linked with each contributing directly or indirectly to a fragile mental healthcare system in Zambia. Figure 3 highlights how these barriers create or perpetuate mental health stigma and the central role of systemic and structural stigma in preventing progress in mental healthcare.

**Table 3:**
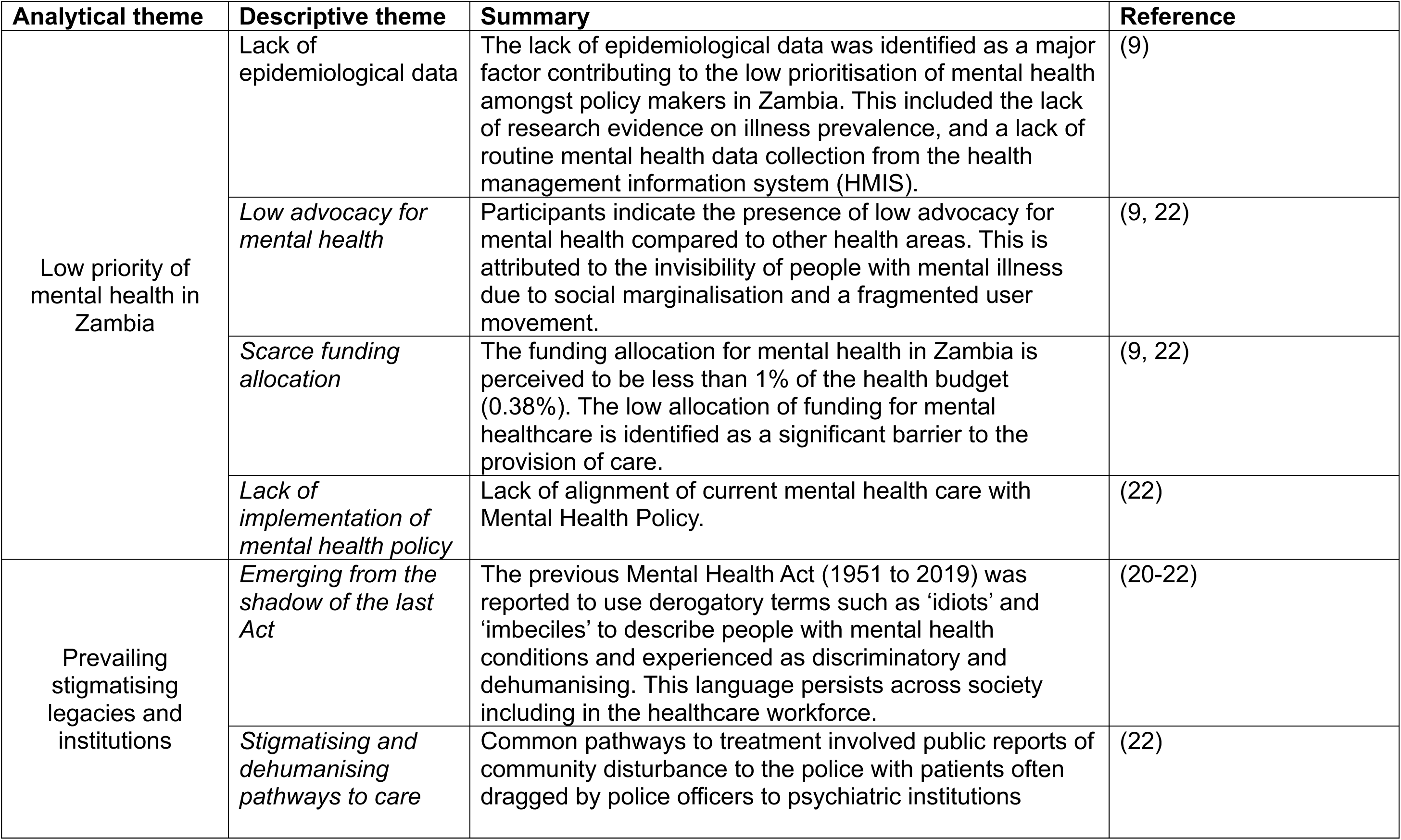
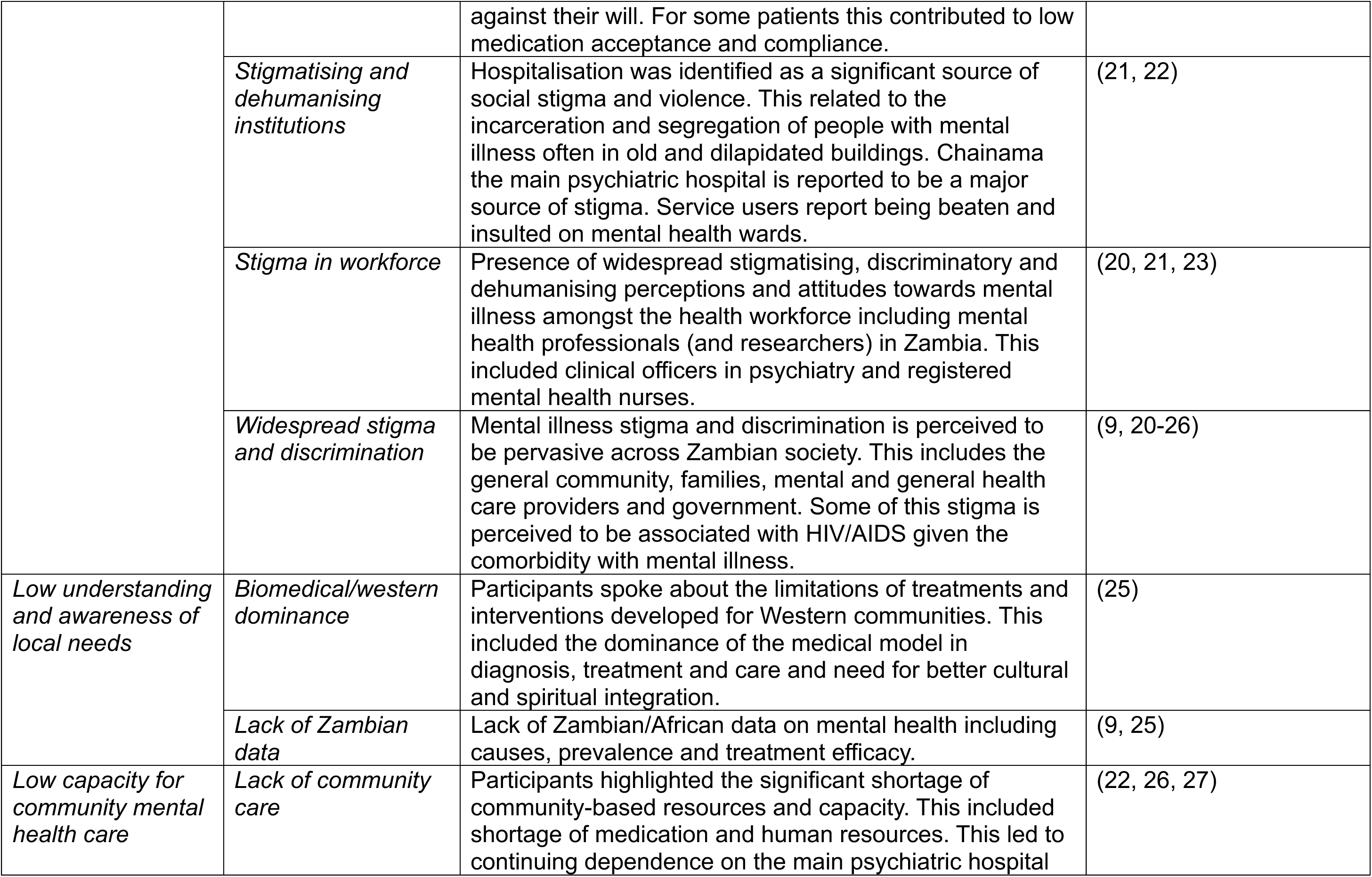
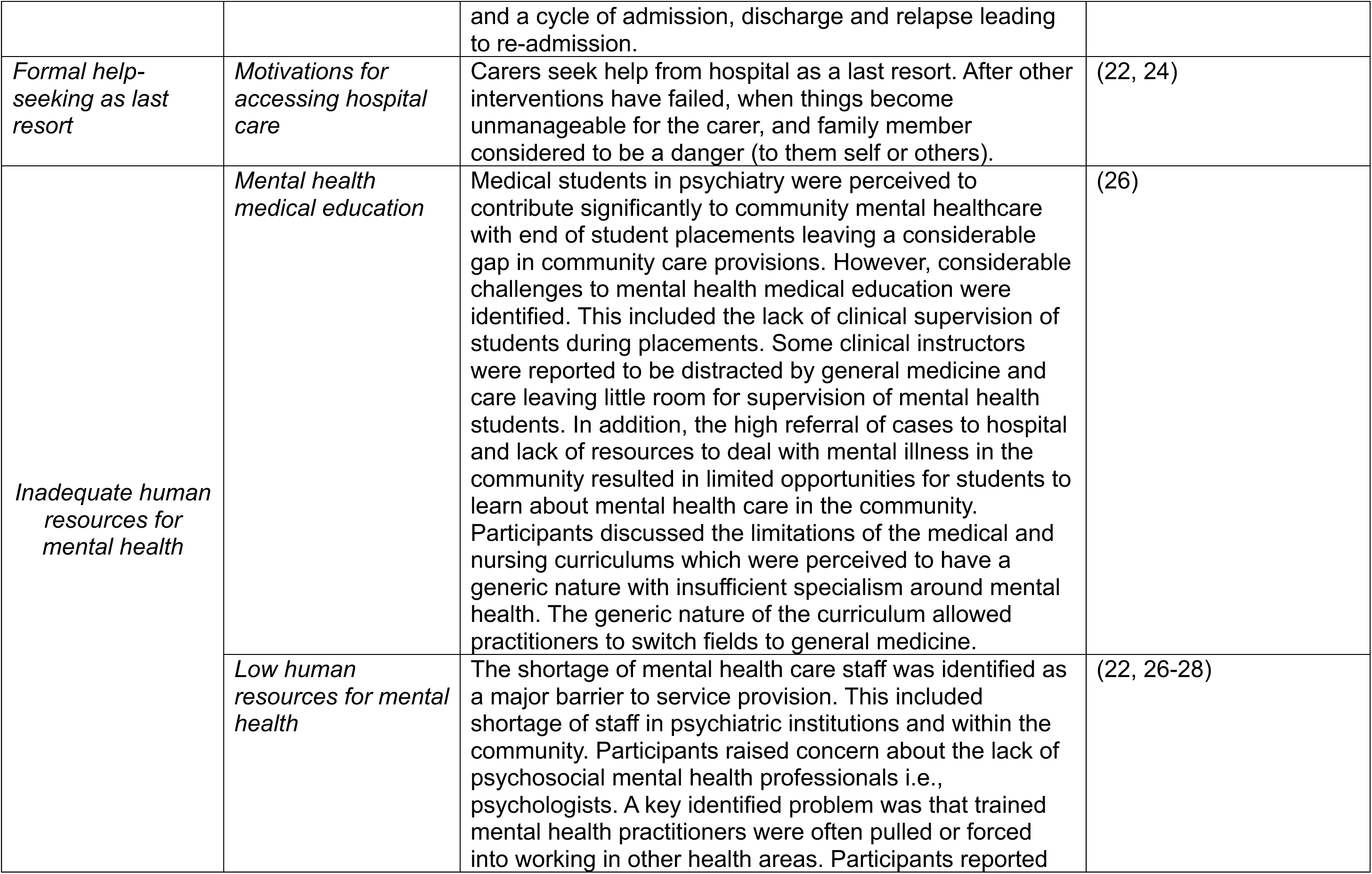
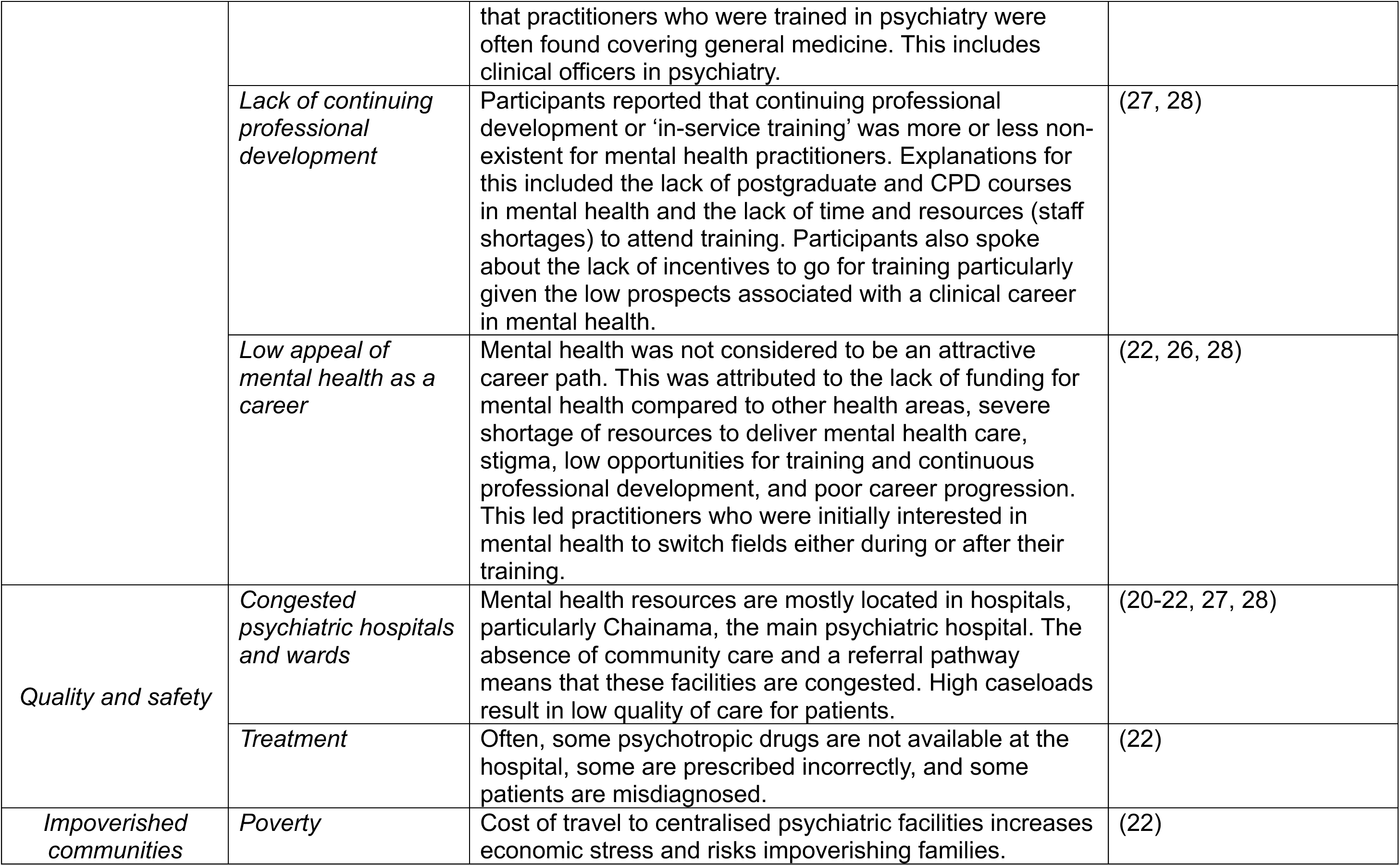
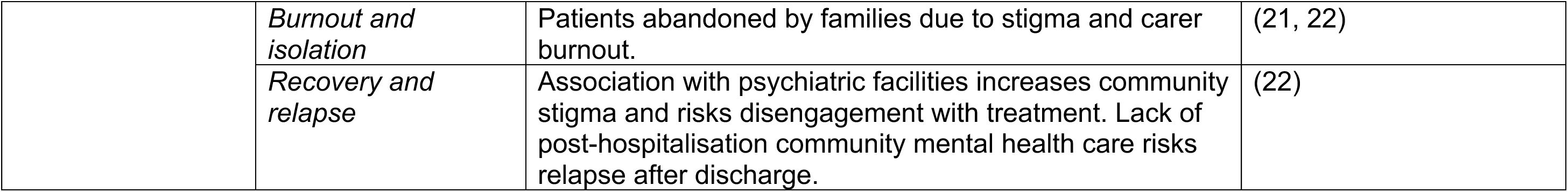
Key themes from thematic synthesis relating to barriers to mental health care in Zambia

**Table 4:** Key themes from thematic synthesis relating to desired solutions and interventions

| **Analytical theme** | **Descriptive theme** | **Summary** | **Reference** |
| --- | --- | --- | --- |
| Strengthen training and education for mental health professionals | Strengthen mental health medical education | Narratives indicated an urgent need to 'motivate and educate' field clinical instructors to meet the syllabus requirements for clinical supervision during student placements. Students would like a more specialised curriculum in mental health. | (26) |
|  | Integrate mental health into entire medical syllabus | Participants emphasised the importance of integrating mental health training with general medicine to better reflect the overlap between mental and physical health and fill the unmet need. | (26) |
|  | Postgraduate courses and training in mental health | Narratives indicated a need for refresher courses/in-service training/postgraduate courses to help update clinical skills and knowledge. | (26) |
|  | Train allied health professionals | Participants emphasised the need to train more allied health professionals, such as psychologists and social workers to allow a multidisciplinary and holistic approach to mental health care. | (28) |
| Build and strengthen community mental health | Quick to use screening tools | Participants felt that easy and quick to use screening tools would help motivate primary care workers to manage mental illness in the community. | (27) |
|  | Buid informal community mental health support | Train informal community mental health workers as a way of filling the gap in community care. Including training faith leaders in basic psychotherapy. | (22) |
| Fill data gap | Increase mental health epidemiology and research and local HMIS data collection | Participants felt that data on the prevalence and impact of mental illness in Zambia was critical to raising awareness of the burden of mental illness to policy makers. This includes the need for better local mental health data collection via HMIS. | (9, 25) |
| Implement multistakeholder approaches to service development | Multisectoral approaches | Participants highlighted the importance of developing and nurturing multi-sectoral partnerships for CAMH (Child and Adolescent Mental Health) advocacy and service delivery. Multi-disciplinary collaborations with mental health professionals such as psychiatrists, psychologists, occupational therapists, and mental health nurses, were regarded as integral to advance CAMH services and promote continuity of care. Additionally, emerging researcher groups and mentorship programmes were viewed as important mechanisms for professional development, in support of global training opportunities, in translating research into effective intervention, and by advancing research through larger multicentre studies. | (25) |
|  | Include families and caregivers | Actively engage family and caregivers in treatment plans | (25) |
|  | Home based interventions | Need for home based interventions that involve school, community and family. | (25) |
|  | Train faith leaders | Train faith leaders to make referrals and do basic psychotherapy | (22) |
|  | Include Lived Experience | Enable full participation of people with Lived Experience of mental illness in decision making | (21) |
| Increase priority of mental health | Integration of mental health with other priority health areas | Increase priority of mental health by teaming up with other priority health areas in Zambia. | (22) |
|  | Awareness and advocacy | Increase awareness and advocacy of mental health using community assistants and wider media. | (22) |
|  | HIV lessons | Draw lessons from HIV to reduce stigma and increase priority of mental health | (9) |
|  | Policy implementation | Strengthen and implement mental health policy | (21) |
|  | Tackle stigma | Tackle mental health stigma in all sectors of society. | (21) |
|  | Increase mental health budget | Increase financial investment in mental health | (28) |
| Digital interventions | Social media and mobile health | Investment in mobile health technologies was identified by a few participants as a promising means to increase community awareness regarding CAMH, while also advancing access to services | (25) |
| African Epistemology | Need for African data | The need for research and adaptation to ensure sociocultural relevance and efficacy of interventions for African communities | (25) |
| Address social determinants and impacts | Address economic stress | Need interventions that address economic stress especially related to travelling for mental health care and having to choose between food and treatment. | (25) |

**Figure 3:**
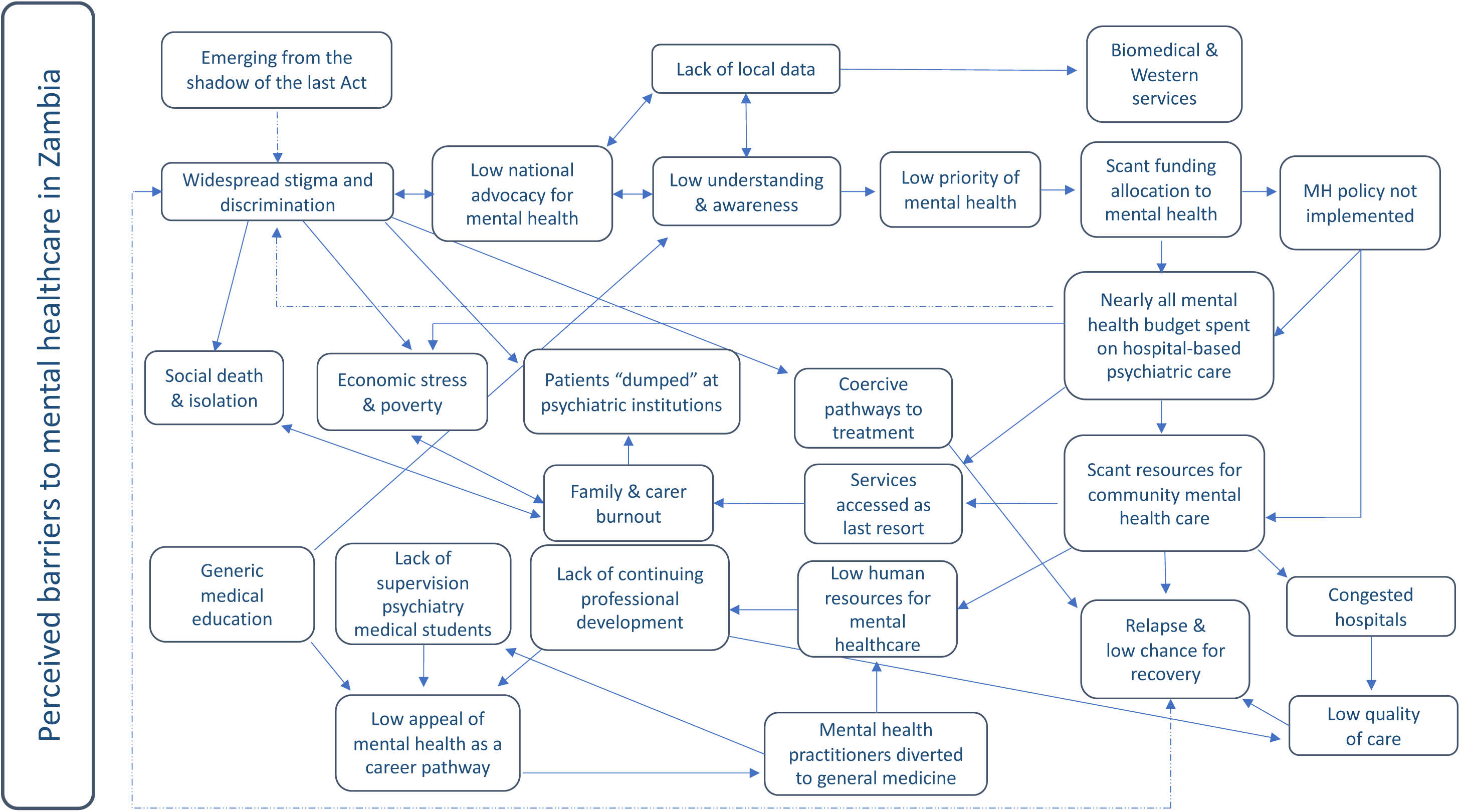
Conceptual map of key barriers to mental health care in Zambia Dashed lines indicate pathways that create or perpetuate mental health s4gma

### Conceptualisation of key themes relating to service barriers (thematic synthesis)

#### Emerging from the shadow of the last Act

The previous Mental Health Act (1951 to 2019) is perceived to be a significant source of mental health stigma. Study participants report that the old Act used discriminating and stigmatising language to describe people with mental illness including referring to them as ‘imbeciles’ and ‘idiots’.(20–22) While the legislation has been revised recently,(2019) participant narratives reveal that the legacy of the last Act remains in the use of discriminating language and the widespread discriminatory attitudes towards people with mental illness.(21) Mental health stigma is perceived to be pervasive and present in all sections of Zambian society, including government and healthcare sectors.(9, 20–26) Service user participants reported experiences of persecution and dehumanisation from their families, community, and health care professionals.(21) Fear of physical violence from “*mental patients*” was perceived to exacerbate negative attitudes and stigma towards people with mental illness in general healthcare. One service user study participants expressed this by saying, “*I can see that they are afraid of me, and fear always makes people behave in negative ways*” (service user study participant).(21)

#### Prevailing stigmatising institutions

Psychiatric institutions are reported to be the main source of formal mental health treatment in Zambia.(21, 22) Chainama, the national psychiatric hospital, seems to be a major source of stigma. Health care provider and service user study participants discussed the lifelong negative consequences of associating with Chainama describing it as a form of social death: “*my future was chopped off*”.(21) Reported negative implications include community rejection of the service user and their family, and unemployment.(21, 22) Participant narratives revealed that psychiatric services were generally accessed as a last resort, when situations became unmanageable for the family, and the affected person is perceived to be a danger.(22, 24) Pathways to psychiatric treatment often include police involvement following reports of community disturbance, where people with mental illness are forcibly taken to psychiatric hospital settings against their will.(22) One service user described experiences of dehumanisation and violence on psychiatric wards “*it’s like they don’t see that you are a person like everybody else, sometimes you are even beaten, we were being beaten and insulted.”*(21) The poor conditions of these facilities, described as old and dilapidated, are perceived to exacerbate the stigma associated with mental illness.(21, 22)

#### Low priority of mental health in Zambia

Mental healthcare is perceived to be severely neglected in Zambia. One study participant, a health district officer, described mental healthcare as the “*Cinderella*” of health services.(21) This neglect is evidenced by the low allocation of funding for mental healthcare (0.38% of health budget), low national advocacy for mental healthcare, lack of local data collection, and poor implementation of the mental health policy.(9, 22) Participants noted that the invisibility of mental illness compared to other health issues contributes to low understanding, awareness, and prioritisation of mental health amongst policy makers.(9) These challenges in policy, funding, advocacy, and data collection appear to be interrelated with each reinforcing the other.(9, 22) Implications include the lack of alignment between current mental healthcare practices and the mental health policy, with a dominance of care based on Western knowledge models.(25) Nearly all mental health resources are located in institutional settings, with little provision for community-based care.(9, 22)

#### Low capacity for community mental healthcare

Participants highlighted a significant shortage of resources and capacity for community-based mental healthcare, including a lack of medication and clinical staff. This shortage perpetuates a dependence on the main psychiatric hospital and a cycle of admission, discharge, relapse and re-admission.(22, 26, 27)

#### Quality and safety

Participants discussed how the absence of community mental healthcare results in congested psychiatric wards. Service provider study participants reported being overwhelmed by high caseloads of patients, with patients often “*dumped*” at the facilities.(22) These high caseloads seem to result in insufficient time to engage with patients. Some patients were perceived to be misdiagnosed or prescribed the wrong medication. Clinical staff also reported frequent shortages of some psychotropic drugs at the hospital.(20–22, 27, 28)

#### Impoverished communities

Reported social consequences of lack of community mental healthcare include delayed help-seeking and treatment provision, poor recovery and social rehabilitation, and relapse.(21, 22) This puts people with mental illness at risk of isolation and abandonment due to caregiver burnout or abandonment by relatives, as a caregiver explained *“Relatives have left us. They get tired early they were many at the beginning but they have fallen off one by one. Because this disease is difficult*”.(21, 22) The lack of visible social recovery may also perpetuate the stigma that people with mental illness are powerless, incapable, and a burden to their families and society.(21) This in turn may contribute to the low advocacy for and prioritisation of mental health.(9) In addition, the cost of travel to centralised facilitates increases economic stress and risks impoverishing families who have to choose between food and treatment.(21, 22) The risk of poverty is perceived to be greater if the affected family member is male and the main or only source of household income.(22)

#### Inadequate human resources for mental health

The lack of resources to provide adequate mental healthcare is described as a significant source of stress for practitioners.(22, 26–28) This includes low medication availability, particularly at the community level, and insufficient human resources to deliver holistic, multidisciplinary mental health care, such as a lack of psychologists and mental health social workers.(22, 26, 28)

Mental health is perceived to be a low-prospect career, affecting both recruitment and retention of the mental health workforce. (22, 26, 28) Clinical educators in mental health discuss student and instructor concerns about the generic nature of the syllabus (for clinical officers in psychiatry and mental health nurses), the lack of specialisation, and the narrow approach to addressing mental illness.(26) In addition, there appears to be inadequate supervision of students during clinical placements, with some reported to be working in isolation.(26) This is attributed to the absence of clinical instructors in psychiatry, who are busy working in general medicine.(26) Participants report that mental health practitioners switch fields due to concerns about low financial and professional prospects or are forced to cover general medicine, with some leaving psychiatry completely as a result.(26) This seems to reduce opportunities for students to gain practical experience in community mental health and create a negative cycle of demotivation about pursuing a career in mental health.(26)

Mental health professionals reported that continuing professional development (CPD) or ‘in-service training’ was more or less non-existent for mental health practitioners.(28) Explanations for this include the lack of postgraduate and CPD courses in mental health, and the lack of time and resources (i.e. staff shortages) to attend training.(27, 28) Participants also spoke about the lack of incentives to go for training, particularly given the low perceived prospects associated with a clinical career in mental health.(27, 28)

### Key themes relating to desired solutions (thematic synthesis)

Key solutions and recommendations from the qualitative literature (Table 4) included increasing the national priority of mental health through awareness-raising and stigma reduction activities, increased financial investment in mental health, and implementation of the mental health policy. Study participants suggested that integrating mental health with high-priority health areas such as maternal health could help overcome some of the funding challenges. Other recommendations included strengthening mental health medical education and postgraduate training, and increasing the mental health knowledge and skills of general medical staff.(26) Primary care workers wanted quick and easy methods to detect mental illness in the community.(27)

The recruitment of community mental health workers and training faith leaders in basic psychotherapy was perceived as valuable for raising awareness of mental health in the community and filling the unmet need.(22) Mental health professionals expressed the need for more psychologists and mental health social workers to allow a multidisciplinary approach to mental health assessment and treatment.(28) Multidisciplinary and multi-stakeholder collaborations between clinicians, communities, schools, families, PWLE of mental illness, and researchers were seen as key to the design of mental health services and interventions in Africa.(21, 25) It was felt that integrating digital technology, such as mobile phones and social media, with mental health interventions could help raise awareness and facilitate access amongst young people.(25)

Participants highlighted the need for local data collection and research, centred around African epistemology, particularly on the incidence and prevalence of common mental disorders in Zambia.(9, 25) Interventions that address the economic impact of mental illness were seen as key for facilitating treatment and recovery.(25)

### Stakeholder feedback and triangulation on scoping review findings

Local stakeholders reported that findings from the thematic synthesis and epidemiological data resonated with their individual and professional experiences of working or living in Zambia and interacting in the mental health space. Mental health professionals were most concerned about the lack of action in operationalising the new Mental Health Act (2019) and in creating policies to align with the Act.

For the local community, the most common concern related to the perceived epidemic of substance use disorders and alcohol dependence across all sectors of society, especially among schoolchildren. NGO stakeholders expressed concern about the lack of multisectoral collaboration and coordination of mental health support in Zambia.

## Discussion

### Summary of key findings

This scoping review found a high prevalence of mental health problems and conditions among adolescents in Zambia, with nearly a third experiencing depressive symptoms, 45.1% reporting problematic drinking, and significant suicidal ideation (31.3%) and behaviour (39.6%). The qualitative analysis highlighted the central role of systemic and structural mental health stigma, rooted in outdated institutional practices and societal attitudes, in preventing the prioritisation of mental health across policy, medical education, and healthcare settings. Reliance on the national psychiatric hospital for mental healthcare and the scarcity of community-based services exacerbates this stigma leading to a cycle of neglect, dehumanisation, and poor recovery for people experiencing mental illness in Zambia.

### Findings in relation to existing literature

To our knowledge, this is the first scoping review of epidemiological and qualitative mental health data in Zambia. Previous reviews of mental health in Sub-Saharan African countries have highlighted poor mental health data coverage. A recent scoping review included surveys from only 12 out of 54 African countries (not including Zambia).(5) Similarly, a review of Sub-Saharan adolescent mental health was based on data from 16 out of 48 Sub-Saharan countries and Zambian data in this review was limited to estimates of suicidal ideation/behaviour.(29) As found in these reviews, comparing estimates across African countries and other world regions is difficult given the significant heterogeneity in social and political contexts, particularly when comparing conflict and post-conflict regions. There is also significant variation in the quantity and quality of reporting and the use of diagnostic and screening tools.(5, 29)

Multi-country studies using standardised methodology, such as school-based mental health surveys GSHS(30) and the WHO STEPS studies,(31) can help provide more comparable prevalence estimates across different regions. Reporting of GSHS data that includes estimates from other Sub-Saharan countries shows that Zambian adolescents have the highest prevalence of depressive symptoms (29.7% in Zambia compared to 22.8% in South Africa),(16) problematic drinking (45.1% Zambia compared to 21.5% in Uganda),(32) and suicidal ideation (Zambia (31.9%), Kenya (27.9%), Botswana (23.1%), Uganda (19.6%) and Tanzania (11.2%)) in these cohorts.(33) These data are corroborated by two large international reviews of the GSHS data that found that Zambia has the highest prevalence of adolescent binge drinking in Africa based on data from 23 African countries, and compared to 72 other low-and middle-income countries (LMICs)(34), and the highest prevalence of youth suicidal ideation based on data from 266,694 school-attending students aged 13–15 years from 49 different LMICs.(30)

For adults, data from the multi-country STEPS study showed similar estimates for suicidal ideation in Zambia and Malawi (7.2 compared to 7.8 in Zambia), but these estimates were twice as high compared to Bhutan (3.1).(31, 35) These data need to be interpreted with caution given that a recent review of global data found significant heterogeneity in estimates of suicidal thoughts and behaviours and suicide in adults within and between LMICs.(36) These differences may reflect the general challenges in psychiatric epidemiology for LMICs relating to quality of surveillance, stigma, disclosure and recording of suicide particularly in relation to criminalisation.(36) Estimates for problematic/harmful alcohol consumption in the general (adult) population in Zambia are difficult to compare with other African countries due to limited data availability from this region and differences in methodology (timeframes, definitions and diagnostic tools).(5) Our estimates for binge (11.6) and harmful drinking (15.3) are similar to the Zambia estimates for heavy episodic drinking reported by the WHO global status report on alcohol and health (11.6 to 13.6) which shows huge variability in estimates across African countries (ranging from 0.6 for Algeria to 63.1 for Equatorial Guinea).(37) Estimates for mental distress are much lower in Zambia than the global prevalence of psychological distress reported by a systematic review in the same year, at 50.0% (2020).(38) However, this review revealed considerable heterogeneity between and within world regions, with no contributing data from Africa for this estimate.(38) In terms of gender differences, our review indicates a higher prevalence of mental distress in women and a higher prevalence of harmful alcohol consumption in men in Zambia. These gender disparities have also been reported in other African countries(5) and is consistent with other international studies but require further corroboration. Our data indicating a higher prevalence of suicidal behaviour in women compared to men also requires further corroboration.(39)

Findings from the qualitative synthesis highlight significant challenges in the current mental healthcare system in Zambia, similar to those experienced in other low-income regions in Sub-Saharan Africa. The availability and quality of care is compromised by the dominance of heavily stigmatised and large psychiatric hospitals that prioritise coercive forms of care as the main source of treatment.(40) Although the WHO guidance to establish community-based mental health care has been adopted as part of national mental health policies, in many Sub-Saharan African countries including Zambia, these have not been fully implemented particularly in relation to the development of robust community alternatives to institutional care and decentralisation of mental healthcare.(41) Our findings identified structural stigma as a major barrier to progress reflected in low prioritisation of mental health in governance including low funding allocation, lack of implementation of mental health policies and legislation, and shortage of medication. Findings from recent evidence reviews reveal a paucity of research on mental health structural stigma in LMICs.(42, 43) An evidence synthesis of structural stigma in Ethiopia identified similar challenges to those identified in our study relating to structural barriers in the scale-up of mental healthcare.(43)

### Implications for policy, practice and research

This review highlights an urgent need to tackle structural stigma across all sectors in Zambian society, including government and healthcare. Addressing this issue is vital to help increase the priority of mental health in Zambia, prevent further discrimination and dehumanisation of people experiencing mental illness, and strengthen the fragile mental healthcare system. Achieving radical shifts in societal attitudes and behaviours requires authentic involvement of PWLE of mental health conditions in decision-making and stigma reduction campaigns.(44) Local stakeholders believe much could be learned from Zambia’s experience of tackling the stigma associated with HIV/AIDS, including the use of peer education programs.(45) Implementing similar strategies could significantly contribute to reducing mental health stigma and improving the overall mental healthcare landscape in Zambia.

All local stakeholders working in governmental mental healthcare in Zambia who provided external feedback on the review emphasized the importance of operationalising the current Mental Health Act (2019) and revising the outdated 2005 mental health policy as key steps to strengthen the current mental health system.

This includes implementation of the National Alcohol Policy (NAP). Despite the adoption of the NAP in 2018, its implementation has been hindered by insufficient funding and lack of government commitment. Stakeholders stressed that ensuring adequate allocation of funding for mental health activities is imperative to translating policies into actionable interventions and improving mental health services nationwide.

Similar to other low-income regions, mental health NGOs in Zambia fill the gap in community mental healthcare in multiple ways, including prevention, awareness-raising and treatment of a wide range of conditions including depression, anxiety, and substance use in children and adults. Their approaches are largely psychosocial, holistic, and non-stigmatising in terms of assessment and treatment. Some, such as StrongMinds, which provides group talk therapy to women and adolescents in urban Zambia, have demonstrated clinical efficacy, with 80% of women free of symptoms six months after intervention.(46, 47) However, NGOs are mostly located in the capital, Lusaka, typically reliant on donor funding, and face challenges integrating their interventions with mainstream mental healthcare services due to the low human resources and capacity in government mental healthcare facilities, exacerbated by the (often undocumented and unofficial) movement of staff to general medicine. Implementing the mental health policy can help protect mental health posts at governmental facilities and provide a budget for integrated community-based mental healthcare activities. This includes building collaborations with NGOs and other community stakeholders (i.e., employment and education sector, traditional healers, community development teams) using asset-based community development and task-shifting approaches.(48) These strategies aim to identify, leverage, and strengthen existing resources in local communities in a low-cost, sustainable and culturally appropriate way. For example, training community health workers, faith leaders, and school teachers in basic psychotherapy can help facilitate prevention, early detection, and support, and help tackle mental health stigma.(48, 49) A key finding from this study is the need to move away from centralised, hospital-based care to community-based territorial care and the need for whole system approaches to integrated care (statutory services working alongside the third sector and local communities) across the whole country.

Our review highlights the need for large-scale population-based studies on mental health conditions in Zambia to help inform priority-setting and policy development. Future research should also focus on the development of locally designed and culturally-tailored ‘Zambia-centric’ solutions for specific issues, such as anti-stigma, suicide prevention (awareness-based), alcohol misuse and addiction in adolescents. Research priority setting and data collection should be guided by a multisectoral board of local stakeholders, including local NGOs and PWLE. Utilising inclusive community participatory methods may help ensure that research is relevant, respectful, and responsive to the needs and contexts of Zambian communities.

### Strengths and limitations

Limitations of the review include the general lack of large-scale epidemiological population studies on the prevalence of mental health conditions in Zambia, the use of self-reported measures by epidemiological studies, and the lack of recent studies (particularly after the COVID-19 pandemic). Strengths of this study include the first comprehensive scoping review of epidemiological and qualitative data in Zambia and triangulation of findings with local stakeholders, which enhances the relevance and applicability of the results.

## Conclusion

This scoping study reveals a high prevalence of mental health problems among adolescents in Zambia, including depressive symptoms, problematic drinking, and suicidal behaviours, which appear to be higher than other Sub-Saharan countries. The study highlights the critical role of systemic and structural mental health stigma in impeding resource allocation and the availability of community-based services.

The reliance on stigmatising and congested psychiatric hospital settings exacerbate these challenges. Addressing these challenges requires a multifaceted approach including early prevention, policy reform and implementation, stigma reduction, deinstutionalisation, and the development of socially and culturally-aligned community-based mental healthcare. Future research should focus on the development of low-cost, culturally-tailored solutions for specific issues like alcohol addiction, stigma reduction, and building a comprehensive mental health database for Zambia.

## Declarations

### Ethics approval and consent to participate

Not applicable – no human participants.

### Consent for publication

Not applicable – no human participants.

### Availability of data and materials

All relevant data are within the manuscript and its supporting files.

### Authors’ contributions

Narinder Bansal – study conceptualisation, data curation, methodology, analysis, investigation, supervision, validation, visualisation, writing – original draft, writing – review and editing.

Petros Andreadis – methodology, analysis, investigation, validation, writing – original draft, writing – review and editing.

Philip Chimponda – validation, writing – review and editing Sandra Barteit - writing – review and editing

Sashi P Sashidharan - review and editing

Ravi Paul – validation, writing – review and editing

## Data Availability

All data produced in the present work are contained in the manuscript

## Acknowledgements

Michael Hobbins

Stakeholders in advisory group (anonymous)

## Abbreviations

PWLE: People with lived experience
PRISMA-ScR: Preferred Reporting Items for Systematic Reviews and Meta-Analyses for Scoping Reviews
GSHS: Global School-Based Student Health Survey of students World Health Organization (WHO)
STEPS: STEPwise approach to non-communicable diseases risk factor surveillance
NGO: Non-governmental organisations
CPD: Continuing Professional Development
LMICs: Low-and Middle-Income Countries
NAP: National Alcohol Policy
SRQ: Self Reporting Questionnaire
NR: Not reported
AUDIT-C: Alcohol Use Disorders Identification Test-Consumption

**Appendix 1:**
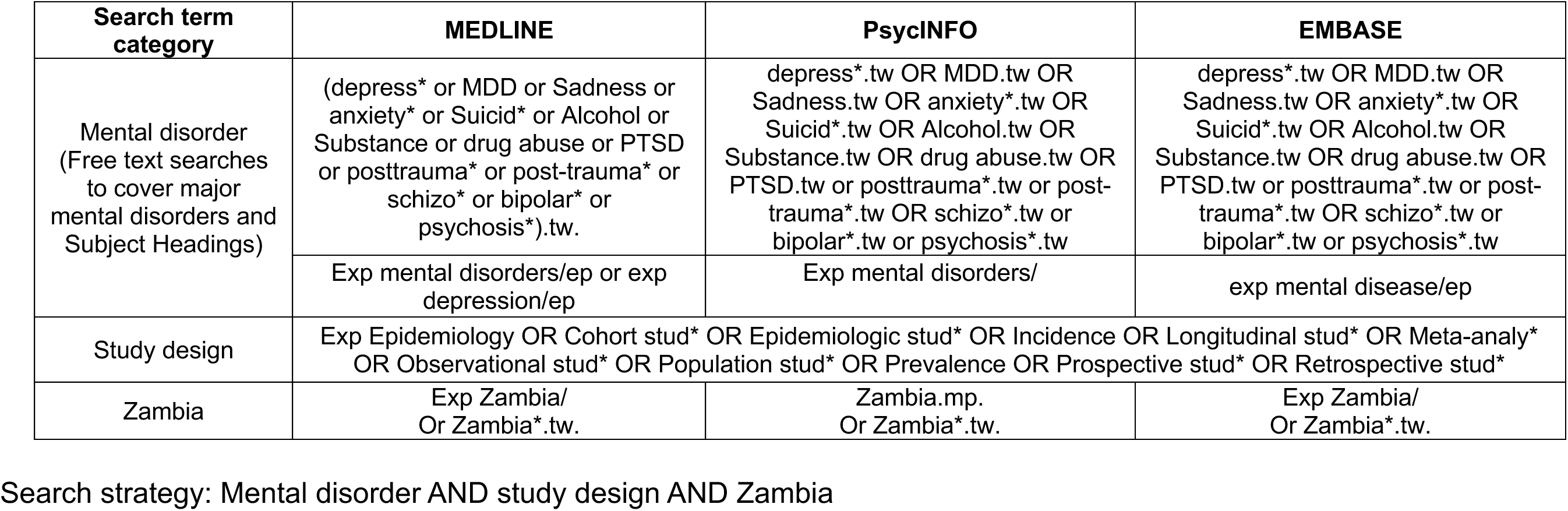
Search strategy (epidemiological literature)

**Appendix 2:**
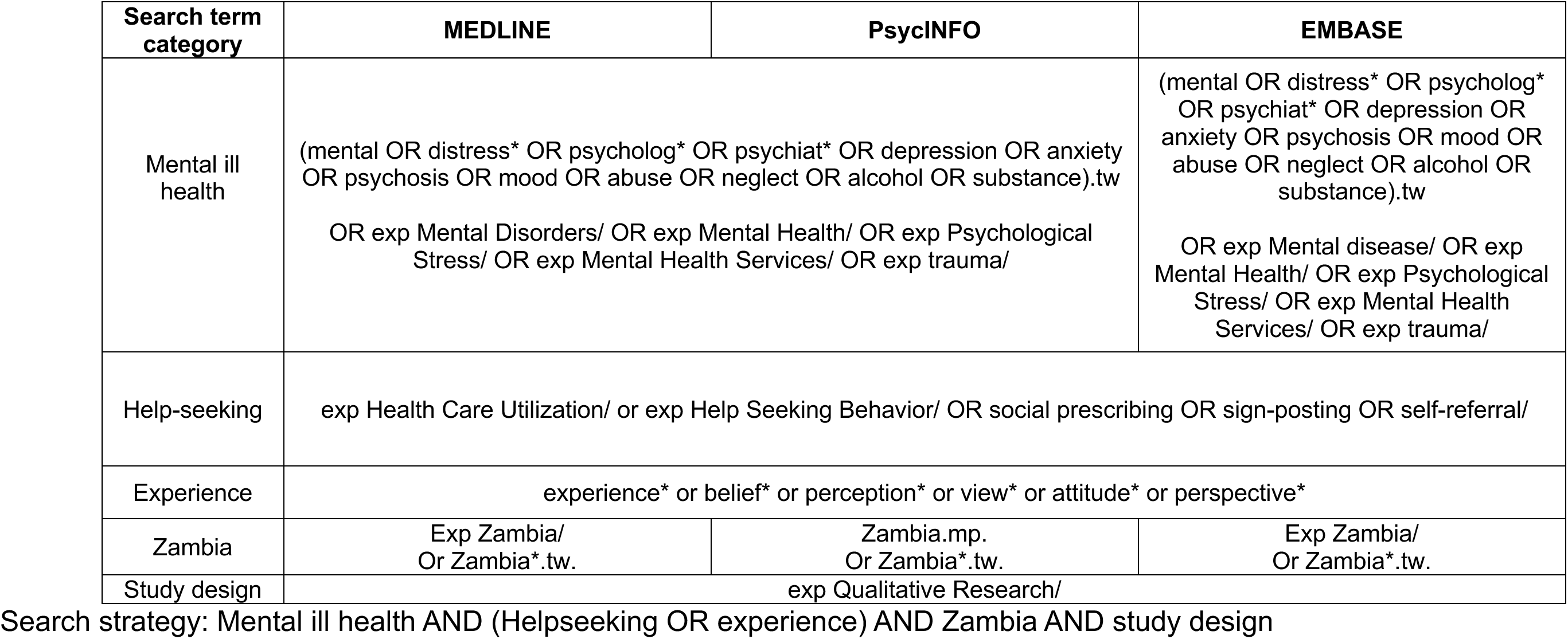
Search strategy (qualitative literature)

